# Exploring drugs and vaccines associated with altered risks and severity of COVID-19: a UK Biobank cohort study of all ATC level-4 drug categories

**DOI:** 10.1101/2020.12.05.20244426

**Authors:** Yong Xiang, Kenneth C.Y. Wong, SO Hon-Cheong

**Affiliations:** School of Biomedical Sciences, Faculty of Medicine, The Chinese University of Hong Kong, Shatin, Hong Kong; CUHK Shenzhen Research Institute, Shenzhen, China; KIZ-CUHK Joint Laboratory of Bioresources and Molecular Research of Common Diseases, Kunming Institute of Zoology and The Chinese University of Hong Kong, China; Department of Psychiatry, Faculty of Medicine, The Chinese University of Hong Kong, Shatin, Hong Kong; Margaret K.L. Cheung Research Centre for Management of Parkinsonism, The Chinese University of Hong Kong, Shatin, Hong Kong; Brain and Mind Institute, The Chinese University of Hong Kong, Shatin, Hong Kong; Hong Kong Branch of the Chinese Academy of Sciences Center for Excellence in Animal Evolution and Genetics, The Chinese University of Hong Kong, Hong Kong SAR, China

**Author notes:** **Correspondence to: Hon-Cheong So**, Lo Kwee-Seong Integrated Biomedical Sciences Building, The Chinese University of Hong Kong, Shatin, Hong Kong.

## Abstract

**Background:** COVID-19 is a major public health concern, yet its risk factors are not well-understood and effective therapies are lacking. It remains unclear how different drugs may increase or decrease the risks of infection and severity of disease.

**Methods:** We studied associations of prior use of all level-4 ATC drug categories (including vaccines) with COVID-19 diagnosis and outcome, based on a prospective cohort of UK Biobank(UKBB). Drug history was based on general practitioner(GP) records. Effects of prescribed medications/vaccinations on the risk of infection, severity of disease and mortality were investigated separately. Hospitalized and fatal cases were categorized as ‘severe’ infection. We also considered different study designs and conducted analyses within infected patients, tested subjects and the whole population respectively, and for 5 different time-windows of prescriptions. Missing data were accounted for by multiple imputation and inverse probability weighting was employed to reduce testing bias. Multivariable logistic regression was conducted which controls for main confounders.

**Results:** We placed a greater focus on protective associations here, as (residual) confounding by indication and comorbidities tends to bias towards harmful effects. Across all categories, statins showed the strongest and most consistent protective associations. Significant protective effects against severe infection were seen among infected subjects (OR for prescriptions within a 12-month window, same below: 0.50, 95% CI:0.42-0.60), tested subjects (OR=0.63, 0.54-0.73) or in the general population (OR=0.49, 0.42-0.57). A number of top-listed drugs with protective effects were also cardiovascular medications, such as angiotensin converting enzyme inhibitors, angiotensin receptor blockers, calcium channel blocker and beta-blockers. Some other drugs showing protective associations included biguanides (metformin), estrogens, thyroid hormones and proton pump inhibitors, among others.

Interestingly, we also observed protective associations by numerous vaccines. The most consistent association was observed for influenza vaccines, which showed reduced odds of infection (OR= 0.73 for vaccination in past year, CI 0.65-0.83) when compared cases to general population controls or test-negative controls (OR=0.60, 0.53-0.68). Protective associations were also observed when severe or fatal infection was considered as the outcome. Pneumococcal, tetanus, typhoid and combined bacterial and viral vaccines (ATC code J07CA) were also associated with lower odds of infection/severity.

Further subgroup and interaction analyses revealed difference in protective effects in different clinical subgroups. For example, protective effects of flu and pneumococcal vaccines were weaker in obese individuals, while we observed stronger protective effects of statins in those with cardiometabolic disorders, such as diabetes, coronary artery disease, hypertension and obesity.

**Conclusions:** A number of drugs, including many for cardiometabolic disorders, may be associated with lower odds of infection/severity of infection. Several existing vaccines, especially flu vaccines, may be beneficial against COVID-19 as well. However, causal relationship cannot be established due to risk of confounding. While further studies are required to validate the findings, this work provides a useful reference for future meta-analyses, clinical trials or experimental studies.

## Introduction

Coronavirus Disease 2019 (COVID-19) has resulted in a pandemic affecting more than a hundred countries worldwide ^1-3^. More than 65 million confirmed infections and 1.5 million fatalities have been reported worldwide as at 20^th^ Nov 2020 (https://coronavirus.jhu.edu/map.html). It is of urgent public interest to gain deeper understanding into the disease, including identifying risk factors (RFs) for infection and severe disease, and uncovering new treatment strategies.

A number of clinical risk factors (e.g. age, obesity, cardiometabolic disorders, renal diseases, multi-comorbidities) ^4-8^ have been suggested to increase the risk to infection or lead to greater risks of complications. However, it is less well-known how different drugs may increase or reduce the risks of COVID-19 or its severity. Drugs with protective effects may be potentially repurposed for the prevention or treatment of the disease. Development of a new drug is often extremely lengthy and costly, while existing drugs with known safety profiles can be brought into practice in a much shorter time-frame.

Here we performed a comprehensive study on all ATC (Anatomical Therapeutic Chemical Classification System) level-4 drug categories (*N*=819) and assessed their associations with susceptibility to and severity of COVID-19 infection in the UK Biobank (UKBB), controlling for possible confounding factors. Vaccines were also included. To our knowledge, this is the most comprehensive analysis of drug associations with COVID-19 to date. While pharmaco-epidemiology studies are typically focused on one or a few drugs, COVID-19 is a new disease and we still have very limited understanding of its pathophysiology and treatment. As a result, a hypothesis-driven approach may have important limitations of missing potential drug associations. In the field of genetic epidemiology, it has been observed that hypothesis-driven candidate gene studies are not as reliable as genome-wide association studies (GWAS) ^9^ which is relatively unbiased, indicating merits of the latter approach. In the same vein, here we adopted a ‘drug-wide’ association study approach, which provides a systematic and unbiased assessment of drug associations. In the present work, we performed rigorous analyses on the impact of medications/vaccinations on the risk of infection, disease severity and mortality. Analyses were also conducted within infected patients, tested subjects and the whole population respectively, and for five different time-windows of prescriptions.

## Methods

### UK Biobank data

The UK Biobank is a large-scale prospective cohort comprising over 500,000 subjects aged 40–69 years who were recruited in 2006–2010 ^10^. In this study, subjects with recorded mortality before 31 Jan 2020 (*N* = 28,930) were excluded, since it was the date for the first recorded case in UK. This study was conducted under project 28732.

### COVID-19 phenotypes

COVID-19 *outcome* data were downloaded from UKBB data portal. Information regarding COVID-19 data in the UKBB was given at http://biobank.ndph.ox.ac.uk/showcase/exinfo.cgi?src=COVID19. Briefly, the latest COVID test results were downloaded on 6 Nov 2020 (last update 3 Nov 2020). We consider inpatient (hospitalization) status at testing as a proxy for severity. Data on date and cause of mortality were also extracted (latest update on 21 Oct 2020). Cases indicated by U07.1 were considered to be (laboratory-confirmed) COVID-19-related fatalities.

A case was considered as having ‘severe COVID-19’ if the subject was hospitalized and/or if the cause of mortality was U07.1. We required both test result and origin to be 1 (positive test and inpatient origin) to be considered as a hospitalized case. For a small number of subjects with initial outpatient origin and positive test result, but changed to inpatient origin and negative result within 2 weeks, we still considered these subjects inpatient cases (i.e. assume the hospitalization was related to the infection).

For a minority of subjects (*N*=19) whose mortality cause was U07.1 but test result(s) was negative within one week, to be conservative, they were excluded from subsequent analyses.

### Medication data

Medication data was obtained from the Primary Care data for COVID-19 research in UKBB (details available at https://biobank.ndph.ox.ac.uk/showcase/showcase/docs/gp4covid19.pdf). In the UK, patients seeking medical advice usually visit a general practitioner (GP) first. Many illnesses are managed under a primary care setting, while most secondary care medical encounters are also reported back to the GP and recorded in their electronic records. We made use of the latest release of GP records released by UKBB, which contains prescription data from two EHR systems (TPP or EMIS) for ∼397,000 UKBB participants. The drug code and issue date of each drug are available.

#### Time window of prescriptions

Since the GP records cover up to ∼50 years’ of prescriptions, we set time windows to restrict prescriptions with a certain time period as the ‘exposure’. The ‘index date’ was defined as (1) the date of the first positive COVID-19 test for infected subjects (for U07.1 cases, the mortality date was regarded as the index date if no test record was found); (2) the date of last test for those tested negative; (3) 3 Nov 2020 for those who were untested.

The issue date of each prescription was available but the duration was not. Time windows were determined by whether the drug was issued within a specified period before the index date. The following windows were considered for medications: 6 months, 1 year, 2 years and 5 years. Narrower time windows (<6 months) may not be desirable and may lead to many prescriptions being missed as the latest issue date was 25 July 2020, but the latest index date was 3 Nov 2020.

As for vaccines, unlike many medications, vaccines are not prescribed regularly and most vaccines only need to be given once or less than a few times; hence a narrow time window is not optimal due to sparsity of data. For seasonal vaccines, namely flu vaccines, they are usually given in autumn (Sep to Nov) or early winter in the UK. A time-window of 6 months will lead to missing most of the flu vaccines given. On the other hand, it is also reasonable to consider a longer time window (e.g. 10 years) as vaccine effects can be more long-lasting ^11^. In view of the above, we considered time windows of 1, 2, 5 and 10 years for vaccinations. For flu vaccines, we defined ‘past 1 year’ as prescriptions from 1^st^ Sep 2019 onwards (and similarly for past *k* years) to account for seasonal nature of vaccination.

#### Mapping to ATC

All the medications were mapped to ATC Classification (https://www.genome.jp/kegg-bin/get_htext?br08303). Drug categories were defined by the 4^th^ level of ATC classification.

### Covariate data

We performed multivariable regression analysis with adjustment for potential confounders including basic demographic variables (age, sex, ethnic group), comorbidities (coronary artery disease[CAD], diabetes[DM], hypertension, asthma, COPD, depression, dementia, history of cancer), blood measurement (e.g. blood urea and creatinine reflecting renal function), indicators of general health (number of medications taken, number of non-cancer illnesses), anthropometric measures (body mass index [BMI]), socioeconomic status (Townsend Deprivation index) and lifestyle risk factor (smoking status). For disease traits, we included information from ICD-10 diagnoses (code 41270), self-reported illnesses (code 20002) and incorporated data from all waves of follow-ups. Subjects with no records of the relevant disease from either self-report or ICD-10 were regarded as having no history of the disease.

### Sets of analysis

We performed a total of 8 sets of analysis (Table 1). The impact of prescribed medication/vaccination on the risk of infection (Model E and F), severity of infection (Model A, C and G) and risk of mortality (Model B, D and H) from COVID-19 were investigated separately. Both hospitalized and fatal cases were grouped under the ‘severe’ category.

**Table 1.** The eight sets of analyses based on infected patients (model A, B), tested subjects (modelsF, G, H) and the population (models C, D, E)

| Model | Cohort 1 | Cohort 2 |
| --- | --- | --- |
| A | Hospitalized or fatal infection (U07.1) (Severe) | Non-hospitalized COVID-19 (Mild) |
| B | U07.1 cases | All other COVID-19 cases |
| C | Hospitalized or fatal infection (U07.1) (Severe) | UKBB subjects without COVID-19 Dx or tested -ve |
| D | U07.1 cases | UKBB subjects without COVID-19 Dx or tested -ve |
| E | Infected | UKBB subjects without COVID-19 Dx or tested -ve |
| F | Infected | Tested -ve |
| G | Hospitalized or fatal infection (U07.1) (Severe) | Tested -ve |
| H | U07.1 cases | Tested -ve |

U07.1 is the code for fatal (laboratory-confirmed) COVID-19 infection based on the latest ICD coding. Dx, diagnosis.

We also considered different study designs and conducted our analyses with different comparison samples. Models A and B are restricted to the infected subjects, while models C, D and E involves comparison of severe, fatal and general infected cases to the general population (with no known diagnosis of COVID-19). On the other hand, models F, G and H compared infected, severe and fatal cases respectively against subjects who were tested negative for SARS-CoV-2.

There were 397,000 subjects in the UKBB with available GP prescription records. Among them, 30,835 subjects have received at least one COVID-19 test, and 3858 were tested positive. There were 1318 cases classified as ‘severe’ (hospitalized or mortality from COVID-19) and 170 fatal cases. In total 393,142 UKBB participants did not have a known diagnosis of COVID-19. The detailed count of participants for each model is listed in Table 2.

**Table 2.** Number of available subjects for analysis for the 8 models

| **Model** | **Cohort 1** | **Cohort 2** | **Total** |
| --- | --- | --- | --- |
| **A** | 1,318 | 2,540 | 3,858 |
| **B** | 170 | 3,688 | 3,858 |
| **C** | 1,318 | 393,142 | 394,460 |
| **D** | 170 | 393,142 | 393,312 |
| **E** | 3,858 | 393,142 | 397,000 |
| **F** | 3,858 | 26,977 | 30,835 |
| **G** | 1,318 | 26,977 | 28,295 |
| **H** | 170 | 26,977 | 27,147 |

Only subjects with available GP prescription records are shown.

### Statistical analysis methods

Logistic regression (using the R package speedglm) was used to examine the impact of medication on different outcomes in the eight sets of analysis. All statistical analyses were conducted using R. The false discovery rate (FDR) approach by Benjamini & Hochberg ^12^ was performed to control for multiple testing. This approach controls the expected proportion of false positives among the rejected null hypotheses.

### Imputation of missing data

Missing values of remaining features were imputed with the R package missRanger. The program is based on missForest, which is an iterative imputation approach based on random forest (RF). It has been widely used and has been shown to produce low imputation errors and good performance in predictive models ^13^. The program missRanger is largely based on the algorithm of missForest, but uses the R package ‘ranger ^14^’ to build RFs, leading to a large improvement in speed. (We found that other packages such as MICE and missForest are computationally too slow to produce results for the large-scale analyses here). Predictive mean matching (pmm) was also employed to avoid imputation with values not present in the original data and increase variance to more realistic levels for multiple imputation (MI). We followed the default settings with pmm.k = 5 and num.trees = 100. We performed the analyses on multiply imputed datasets (imputed for 10 times) and combined the results by Rubin’s rules^15^ using the mi.meld function under the R package amelia. Another advantage of missRanger is that out-of-bag errors (in terms of classification errors or normalized root-mean-squared error) could be computed which provides an estimate of imputation accuracy.

### Inverse probability weighting of the probability of being tested

Bias due to non-random testing has been discussed previously in other works ^16,17^. As a person has to be tested to be diagnosed of COVID-19, factors leading to increased risk of being tested will also lead to an apparent increase in the risk of infection ^17^. In addition, it has been raised that collider bias can occur when conditioned on the tested group and results in spurious associations, for example between a risk factor and COVID-19 severity if both increases the the probability of being tested [Pr(tested)]. One way to reduce this kind of bias is to employ inverse probability weighting (IPW) of Pr(tested). Essentially, we wish to create a pseudo-population or mimic a scenario under which testing is more *random* instead of selected for certain subgroups. The IPW approach unweighs those who are less likely tested and downweighs those who have a high chance of being tested. This may create more unbiased estimates of the effects of drugs.

We took reference to the approach described in ^16^ to analyze the data with IPW. Following our recent work ^18^ which aims to predict COVID-19 severity with machine learning (ML), here we also employed an ML model (XGboost) to predict Pr(tested) based on a range of factors. An advantage of using ML models is that non-linear and complex interactions can be considered, which may improve predictive performance over logistic models. We employed the same set of predictors as our previous work, and followed the same analysis strategy of hyper-parameter tuning and cross-validation to obtain predicted probabilities (please refer to^18^ for details). Beta-calibration ^19^ was performed and the resulting average AUC was 0.622. The predicted probabilities [i.e. Pr(tested)] were used to construct weights for IPW. Stabilized weights^20^ were used.

### Subgroup analysis

For selected drugs showing tentative protective effects, we also performed further subgroup and interaction analyses. These drugs included cardiovascular medications listed in Table 3, four vaccines with protective associations (influenza, pneumococcal, typhoid and combined bacterial/viral vaccines), and other top drugs with consistent protective associations across multiple models/time-windows as listed in Table 6.

**Table 3.** Cardiometabolic medications showing significant protective associations within time windows of 6, 12 and 24 months

| Window | Model | ATC code | OR | conf.low | conf.high | p | FDR.BH | Full name |
| --- | --- | --- | --- | --- | --- | --- | --- | --- |
| 1yrs | F | A10AE | 0.52 | 0.32 | 0.84 | 6.88E-03 | 9.82E-02 | A10AE Insulins and analogues for injection, long-acting |
| 2yrs | A | A10BA | 0.60 | 0.42 | 0.86 | 5.00E-03 | 8.26E-02 | A10BA Biguanides |
| 1yrs | C | A10BA | 0.67 | 0.51 | 0.88 | 4.01E-03 | 2.24E-02 | A10BA Biguanides |
| 2yrs | C | A10BA | 0.68 | 0.52 | 0.90 | 5.79E-03 | 3.22E-02 | A10BA Biguanides |
| 0.5yr | F | C07AB | 0.78 | 0.68 | 0.89 | 3.56E-04 | 1.28E-02 | C07AB Beta blocking agents, selective |
| 1yrs | F | C07AB | 0.80 | 0.70 | 0.91 | 7.59E-04 | 2.24E-02 | C07AB Beta blocking agents, selective |
| 2yrs | F | C07AB | 0.78 | 0.69 | 0.88 | 9.10E-05 | 3.52E-03 | C07AB Beta blocking agents, selective |
| 1yrs | C | C08CA | 0.76 | 0.64 | 0.90 | 1.31E-03 | 8.59E-03 | C08CA Dihydropyridine derivatives |
| 2yrs | C | C08CA | 0.78 | 0.66 | 0.92 | 3.27E-03 | 2.15E-02 | C08CA Dihydropyridine derivatives |
| 0.5yr | A | C09AA | 0.68 | 0.53 | 0.87 | 2.11E-03 | 2.72E-02 | C09AA ACE inhibitors, plain |
| 1yrs | A | C09AA | 0.68 | 0.54 | 0.86 | 1.15E-03 | 1.87E-02 | C09AA ACE inhibitors, plain |
| 2yrs | A | C09AA | 0.67 | 0.54 | 0.84 | 5.87E-04 | 1.85E-02 | C09AA ACE inhibitors, plain |
| 0.5yr | C | C09AA | 0.75 | 0.62 | 0.91 | 3.15E-03 | 1.23E-02 | C09AA ACE inhibitors, plain |
| 1yrs | C | C09AA | 0.61 | 0.51 | 0.74 | 1.59E-07 | 2.86E-06 | C09AA ACE inhibitors, plain |
| 2yrs | C | C09AA | 0.63 | 0.53 | 0.75 | 2.84E-07 | 5.60E-06 | C09AA ACE inhibitors, plain |
| 1yrs | E | C09AA | 0.79 | 0.72 | 0.88 | 1.40E-05 | 1.67E-04 | C09AA ACE inhibitors, plain |
| 2yrs | E | C09AA | 0.81 | 0.73 | 0.90 | 5.38E-05 | 6.06E-04 | C09AA ACE inhibitors, plain |
| 0.5yr | G | C09AA | 0.68 | 0.56 | 0.83 | 1.13E-04 | 3.18E-03 | C09AA ACE inhibitors, plain |
| 1yrs | G | C09AA | 0.71 | 0.59 | 0.85 | 2.80E-04 | 8.26E-03 | C09AA ACE inhibitors, plain |
| 2yrs | G | C09AA | 0.71 | 0.59 | 0.85 | 1.41E-04 | 5.24E-03 | C09AA ACE inhibitors, plain |
| 1yrs | C | C09CA | 0.68 | 0.54 | 0.85 | 7.58E-04 | 5.35E-03 | C09CA Angiotensin II receptor blockers, plain |
| 2yrs | C | C09CA | 0.73 | 0.58 | 0.90 | 3.97E-03 | 2.41E-02 | C09CA Angiotensin II receptor blockers, plain |
| 0.5yr | G | C09CA | 0.72 | 0.56 | 0.91 | 7.00E-03 | 9.51E-02 | C09CA Angiotensin II receptor blockers, plain |
| 1yrs | G | C09CA | 0.69 | 0.55 | 0.87 | 1.95E-03 | 3.84E-02 | C09CA Angiotensin II receptor blockers, plain |
| 2yrs | G | C09CA | 0.72 | 0.58 | 0.90 | 3.93E-03 | 8.79E-02 | C09CA Angiotensin II receptor blockers, plain |
| 0.5yr | A | C10AA | 0.57 | 0.47 | 0.68 | 3.37E-09 | 1.74E-07 | C10AA HMG CoA reductase inhibitors |
| 1yrs | A | C10AA | 0.50 | 0.42 | 0.60 | 2.87E-13 | 9.36E-11 | C10AA HMG CoA reductase inhibitors |
| 2yrs | A | C10AA | 0.49 | 0.40 | 0.58 | 1.55E-14 | 5.38E-12 | C10AA HMG CoA reductase inhibitors |
| 0.5yr | C | C10AA | 0.79 | 0.68 | 0.91 | 1.20E-03 | 5.19E-03 | C10AA HMG CoA reductase inhibitors |
| 1yrs | C | C10AA | 0.49 | 0.42 | 0.57 | 2.97E-21 | 1.70E-19 | C10AA HMG CoA reductase inhibitors |
| 2yrs | C | C10AA | 0.49 | 0.43 | 0.57 | 7.09E-21 | 5.59E-19 | C10AA HMG CoA reductase inhibitors |
| 1yrs | D | C10AA | 0.50 | 0.34 | 0.74 | 5.28E-04 | 5.91E-03 | C10AA HMG CoA reductase inhibitors |
| 2yrs | D | C10AA | 0.50 | 0.34 | 0.74 | 4.38E-04 | 6.28E-03 | C10AA HMG CoA reductase inhibitors |
| 1yrs | E | C10AA | 0.83 | 0.77 | 0.91 | 1.69E-05 | 1.94E-04 | C10AA HMG CoA reductase inhibitors |
| 2yrs | E | C10AA | 0.86 | 0.79 | 0.93 | 3.09E-04 | 2.71E-03 | C10AA HMG CoA reductase inhibitors |
| 0.5yr | G | C10AA | 0.66 | 0.57 | 0.76 | 2.55E-08 | 1.67E-06 | C10AA HMG CoA reductase inhibitors |
| 1yrs | G | C10AA | 0.63 | 0.54 | 0.73 | 4.15E-10 | 5.71E-08 | C10AA HMG CoA reductase inhibitors |
| 2yrs | G | C10AA | 0.63 | 0.54 | 0.72 | 2.65E-10 | 5.63E-08 | C10AA HMG CoA reductase inhibitors |

Only results with FDR<0.1 are shown.
OR, odds ratio; conf.low, lower 95% CI for OR; conf.high, upper 95% CI for OR; FDR.BH, false discovery rate by the Benjamini Hochberg method.

Subgroup analysis was performed with respect to main demographic features (age, sex and ethnicity) and main comorbidities (same as the diseases listed under ‘covariate data’). We also compared log(OR) (i.e. logistic regression coefficient) estimates across the subgroups with or without the risk factor of interest. The test statistic was obtained by 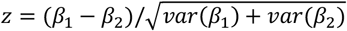, where *β*_1_ and *β*_2_ refer to the coefficients under the 2 independent subgroups. For flu vaccines, the subgroup analysis was restricted within the past 1, 2, 5, and 10 years. For other drugs, the subgroup analysis was restricted to the past 0.5, 1, 2, and 5 years.

### Interaction analysis

As a complementary approach, we also performed analysis with a logistic model including an interaction term (drug*risk_factor). The same set of drugs and risk factors were studied. The two approaches are similar in principle; however stratified analysis yields more unbiased estimates if confounders have subgroup-dependent associations, while the interaction-term approach produces more precise (lower SE) estimates (hence higher power to detect interactions). ^21^

### Controlling for other drugs

We also performed additional regression analyses controlling for other top-ranked drugs. Two sets of analyses were conducted. In the first set of analysis, we controlled for the top 10 or 20 protective and harmful drugs in each time-window and model. As for the second analysis, for drugs with protective associations, we controlled all other protective drugs with FDR<0.05 or 0.1. (This analysis was performed for protective drugs only as there were too many drugs associated with harmful effects to be included as covariates).

## Results

Due to the large number of models and drugs being studied, we shall highlight the main results and findings from different sensitivity analysis.

Confounding by indication and other comorbidities is unavoidable and in particular, drugs showing harmful effects may possibly be explained by such confounding. On the other hand, as it is expected that most diseases tend to *increase* the risk/severity of infection, drugs showing *protective* effects are much less likely to be affected by confounding, and such associations may be relatively more reliable. We therefore place a greater emphasis on protective drugs in the sections below, although main drugs with harmful effects will also be briefly discussed.

A summary of the demographic and covariate data of the original UKBB dataset is shown in Table S1. The missing rates and out-of-bag (OOB) errors for different variables from multiple imputations are shown in Table S2.

### Analysis on subjects with available GP records and multiple imputation of covariates

Full results of all drug categories across all time windows (including 6, 12, 24, 60 and 120 months; the last time-window only for vaccines) are shown in Tables S6 to S10. All protective associations (with at least nominal significance i.e. p<0.05) are shown in Table S3, while all association results with vaccines are presented in Table S4. For drugs associated with increased odds of infection/severity, we also summarized the top 10 drugs (ranked by p-value) from each model and time window, and put them together in Table S5.

#### Overview

Across all categories, statins showed the strongest and most consistent protective associations. Highly significant protective effects were seen across infected subjects, tested or the whole population. The most consistent evidence is for models A, C, D and G, which suggests its effect in reducing the severity or mortality of infection. Albeit with smaller effect sizes, we also observed that statins might be linked to lower susceptibility to infection (model E). Interestingly, a number of top-listed drugs are also cardiovascular medications, such as angiotensin converting enzyme inhibitors (ACEI), angiotensin receptor blockers (ARB), calcium channel blocker (CCB) and beta-blockers.

For simplicity, odds ratios (OR) are presented for a time horizon of 1 year if not further specified.

#### Drugs for cardiometabolic disorders

Significant protective associations (FDR<0.1) are shown in Table 3. Statins showed protective effects across models A, C, D, E and G. Significant protective effects against severe infection were seen among infected subjects (OR for prescriptions within a 12-month window, same below: 0.50, 95% CI:0.42-0.60), tested subjects (OR=0.63, 0.54-0.73) or when comparing severe cases to the general population (OR=0.49, 0.42-0.57). In addition, protective association against fatality was observed (1-year OR: 0.51, CI 0.34 – 0.74). Statins was also associated with lower susceptibility to infection, with OR of 0.83 (CI: 0.77 – 0.91) and 0.86 (CI: 0.79 – 0.93) for prescriptions within 1 year and 2 years respectively.

Another group of drugs with highly consistent protective associations are *ACEI and ARB*. ACEI showed protective associations against severe disease among infected subjects (model A: OR for 1-year time window=0.68, CI: 0.54 – 0.86), and when comparing to the general population (model C: OR 1-year=0.61, CI: 0.51 – 0.74) or test-negative subjects (model G: OR 1-year=0.71, CI: 0.59 – 0.85). We also observed association with lower odds of infection at a population level (model E: OR 1-year=0.81, CI: 0.73 – 0.90); the effect size seemed to decrease over longer time windows. ARBs also showed protective associations against severe disease in the population (model C: OR 1-year=0.54, CI: 0.54-0.85) or among tested individuals (model G: OR 1-year = 0.68, CI: 0.55 – 0.87).

Biguanides (mainly metformin) were associated with lower odds of severe illness among the infected (model A: OR for 2-year time window= 0.60, CI: 0.42-0.86) and in the population (model C; OR 1-year=0.67, CI: 0.51 – 0.88). Other drugs of interest include beta-blockers which were associated with lower risk of infection when comparing test-positive vs test-negative subjects (model F, OR 1-year=0.80, CI: 0.70-0.91), and CCBs (C08CA) which were associated with lower odds of severe disease in the population (model C, OR 1-year: 0.76, CI: 0.64 – 0.90).

#### Vaccines

Significant associations for vaccines (FDR<0.1) are shown in Table 4. As for vaccines, one of the most consistent associations was observed for influenza vaccines. Protective associations were observed across models B to H, and across all time windows. Flu vaccination was associated with lower odds of infection when compared to population controls (model E; OR 1-year=0.73, CI: 0.65 – 0.83) or compared to test-negative individuals (model F; OR 1-year=0.60, CI: 0.53 – 0.68). Similar protective effects were also observed when restricting the cases to severe cases (model C: OR 1-year=0.74; CI: 0.60-0.91; model G: OR 1-year=0.61, CI: 0.50 – 0.76). Association with lower odds of mortality was also observed, although the confidence interval is wide as the number of fatal cases was small (model D: OR 1-year=0.28, CI: 0.13-0.63; model H: OR 1-year=0.23, CI: 0.11 – 0.52). The effect sizes in general became weaker with longer time windows.

**Table 4.** Vaccines with significant protective associations (FDR<0.1) within time windows of 1, 2, 5 and 10 years

| Window | Model | ATC code | OR | conf.low | conf.high | p | FDR.BH | Name of vaccine |
| --- | --- | --- | --- | --- | --- | --- | --- | --- |
| 5yrs | E | J07AL | 0.70 | 0.55 | 0.89 | 3.81E-03 | 2.67E-02 | J07AL Pneumococcal vaccines |
| 10yrs | E | J07AL | 0.78 | 0.67 | 0.91 | 1.89E-03 | 1.32E-02 | J07AL Pneumococcal vaccines |
| 1yrs | F | J07AL | 0.50 | 0.31 | 0.82 | 5.29E-03 | 8.11E-02 | J07AL Pneumococcal vaccines |
| 2yrs | F | J07AL | 0.59 | 0.42 | 0.82 | 1.59E-03 | 3.56E-02 | J07AL Pneumococcal vaccines |
| 5yrs | F | J07AL | 0.61 | 0.47 | 0.79 | 1.47E-04 | 5.18E-03 | J07AL Pneumococcal vaccines |
| 10yrs | F | J07AL | 0.67 | 0.57 | 0.78 | 9.39E-07 | 6.57E-06 | J07AL Pneumococcal vaccines |
| 10yrs | G | J07AL | 0.67 | 0.51 | 0.87 | 3.32E-03 | 1.71E-02 | J07AL Pneumococcal vaccines |
| 10yrs | E | J07AM | 0.65 | 0.45 | 0.92 | 1.60E-02 | 6.58E-02 | J07AM Tetanus vaccines |
| 5yrs | F | J07AM | 0.45 | 0.29 | 0.68 | 1.94E-04 | 5.92E-03 | J07AM Tetanus vaccines |
| 10yrs | F | J07AM | 0.49 | 0.34 | 0.71 | 1.69E-04 | 5.92E-04 | J07AM Tetanus vaccines |
| 10yrs | E | J07AP | 0.86 | 0.76 | 0.97 | 1.88E-02 | 6.58E-02 | J07AP Typhoid vaccines |
| 5yrs | F | J07AP | 0.70 | 0.58 | 0.84 | 1.60E-04 | 5.23E-03 | J07AP Typhoid vaccines |
| 10yrs | F | J07AP | 0.76 | 0.67 | 0.88 | 1.18E-04 | 5.51E-04 | J07AP Typhoid vaccines |
| 10yrs | G | J07AP | 0.74 | 0.58 | 0.95 | 1.61E-02 | 5.23E-02 | J07AP Typhoid vaccines |
| 1yrs | C | J07BB | 0.74 | 0.60 | 0.91 | 3.80E-03 | 4.94E-02 | J07BB Influenza vaccines |
| 2yrs | C | J07BB | 0.75 | 0.62 | 0.90 | 2.02E-03 | 1.45E-02 | J07BB Influenza vaccines |
| 1yrs | D | J07BB | 0.28 | 0.13 | 0.63 | 1.92E-03 | 2.50E-02 | J07BB Influenza vaccines |
| 2yrs | D | J07BB | 0.30 | 0.15 | 0.60 | 7.22E-04 | 9.07E-03 | J07BB Influenza vaccines |
| 1yrs | E | J07BB | 0.73 | 0.65 | 0.83 | 5.93E-07 | 7.71E-06 | J07BB Influenza vaccines |
| 2yrs | E | J07BB | 0.75 | 0.68 | 0.84 | 4.83E-07 | 8.79E-06 | J07BB Influenza vaccines |
| 5yrs | E | J07BB | 0.80 | 0.73 | 0.88 | 7.01E-06 | 9.95E-05 | J07BB Influenza vaccines |
| 10yrs | E | J07BB | 0.82 | 0.75 | 0.89 | 6.70E-06 | 9.38E-05 | J07BB Influenza vaccines |
| 1yrs | F | J07BB | 0.60 | 0.53 | 0.68 | 2.94E-15 | 3.23E-14 | J07BB Influenza vaccines |
| 2yrs | F | J07BB | 0.62 | 0.55 | 0.70 | 4.38E-16 | 1.87E-13 | J07BB Influenza vaccines |
| 5yrs | F | J07BB | 0.66 | 0.60 | 0.73 | 7.67E-16 | 1.76E-13 | J07BB Influenza vaccines |
| 10yrs | F | J07BB | 0.67 | 0.61 | 0.74 | 5.16E-17 | 7.22E-16 | J07BB Influenza vaccines |
| 1yrs | G | J07BB | 0.61 | 0.50 | 0.76 | 4.35E-06 | 4.78E-05 | J07BB Influenza vaccines |
| 2yrs | G | J07BB | 0.62 | 0.52 | 0.75 | 8.86E-07 | 5.38E-05 | J07BB Influenza vaccines |
| 5yrs | G | J07BB | 0.69 | 0.59 | 0.81 | 8.14E-06 | 5.31E-04 | J07BB Influenza vaccines |
| 10yrs | G | J07BB | 0.69 | 0.59 | 0.80 | 9.82E-07 | 1.28E-05 | J07BB Influenza vaccines |
| 1yrs | H | J07BB | 0.23 | 0.11 | 0.52 | 4.04E-04 | 4.44E-03 | J07BB Influenza vaccines |
| 2yrs | H | J07BB | 0.25 | 0.12 | 0.50 | 9.64E-05 | 3.41E-03 | J07BB Influenza vaccines |
| 5yrs | H | J07BB | 0.44 | 0.27 | 0.72 | 1.13E-03 | 3.40E-02 | J07BB Influenza vaccines |
| 10yrs | H | J07BB | 0.50 | 0.32 | 0.76 | 1.44E-03 | 1.87E-02 | J07BB Influenza vaccines |
| 10yrs | F | J07BC | 0.86 | 0.75 | 0.99 | 3.67E-02 | 8.56E-02 | J07BC Hepatitis vaccines |
| 10yrs | E | J07CA | 0.91 | 0.84 | 0.99 | 2.90E-02 | 8.12E-02 | J07CA Bacterial and viral vaccines, combined |
| 1yrs | F | J07CA | 0.56 | 0.38 | 0.84 | 4.30E-03 | 6.94E-02 | J07CA Bacterial and viral vaccines, combined |
| 2yrs | F | J07CA | 0.71 | 0.57 | 0.89 | 3.05E-03 | 5.89E-02 | J07CA Bacterial and viral vaccines, combined |
| 10yrs | F | J07CA | 0.85 | 0.78 | 0.94 | 7.85E-04 | 2.20E-03 | J07CA Bacterial and viral vaccines, combined |
| 10yrs | G | J07CA | 0.78 | 0.66 | 0.92 | 3.94E-03 | 1.71E-02 | J07CA Bacterial and viral vaccines, combined |

In view of the significant findings, we repeated analyses on flu vaccines with other approaches to define the exposure (Table S18). First we defined the exposure based on the *season* of vaccination instead of any vaccines received in the past *k* years. For people who have received flu vaccination in 2019-20 (regardless of vaccination in other years), the OR for infection was 0.60 (CI 0.53-0.68) compared to those who have not (test-negative subjects as controls, model F). The OR was attenuated to 0.76 (OR 0.67– 0.87) if the exposure was defined as flu vaccination in 2015-2016 (regardless of vaccination in other years).

We then narrowed down the exposure as receiving flu vaccine in the last season (2019-20) but *not* in 2018-19; the resulting OR was 0.67 (CI 0.53-0.83). On the other hand, if we considered exposure as vaccination in 18-19 but not 19-20, the OR became weaker and non-significant (OR=0.80, 0.63-1.01). Interestingly, those who received the vaccine *consecutively* for the last 2 seasons appeared to have slightly stronger protection from infection (OR=0.59, 0.51-0.69). A similar pattern of association was observed for model E (population controls). In general, more recent vaccination was associated with stronger protective effects.

Pneumococcal vaccines were also associated with protective effects, especially when comparing within tested subjects (model F: OR 1-year=0.50, CI:0.31-0.82), which shows a trend of attenuation with longer time windows (OR for 10-year window=0.67, CI: 0.51– 0.87). Another group of vaccines showing protective effects is J07CA (bacterial and viral vaccines) which was significant under model F (OR for 1-year window: 0.56, CI: 0.38 – 0.84); it also showed weakening of effect over time. Other significant associations included tetanus and typhoid vaccines which were observed to be protective against infections.

#### Other drugs showing protective associations

Significant results for other drugs having protective effects and FDR<0.1 are shown in Table 5. As for other drugs, proton pump inhibitors (PPI) was associated with lower odds of infection when we compared test-positive against test-negative patients (model F: OR 1 year=0.77, CI: 0.71 – 0.83); the OR showed a gradient with largest effect within 6 month of use (OR=0.72) and became weaker at 5-year time window (OR=0.87). PPI was also significantly associated with lower severity of disease. Estrogens (ATC G03CA) was consistently linked to lower risk of infection and severity in the tested population (model F: 1-year OR 0.67, CI: 0.58 – 0.78) which showed attenuation of effect over time. The greatest effect size was noted within 6 month of use (OR=0.63) which was attenuated for a 5-year time window (OR=0.73). Similar protective associations were observed for model G with severity as the outcome. Prior use of thyroid hormones was consistently associated with lower risk of infection and severity, no matter the general population or test-negative individuals were considered as controls. The ORs were similar across all time windows. For model E (infected vs population), OR for 1 year time window was 0.80 (CI 0.71 to 0.92), which was close to the effect size under model F (infected vs test-negative). For model C (hospitalized/fatal cases vs population), OR for 1 year time window was 0.62 (CI 0.48 to 0.79) and estimates were similar under model G.

**Table 5.** Other drugs with significant protective associations (FDR<0.1) within time windows of 6, 12 and 24 months

| Window | Model | ATC code | OR | conf.low | conf.high | p | FDR.BH | Full name |
| --- | --- | --- | --- | --- | --- | --- | --- | --- |
| 1yrs | A | A02BC | 0.77 | 0.65 | 0.91 | 2.37E-03 | 3.22E-02 | A02BC Proton pump inhibitors |
| 2yrs | A | A02BC | 0.77 | 0.66 | 0.90 | 1.05E-03 | 3.04E-02 | A02BC Proton pump inhibitors |
| 0.5yr | F | A02BC | 0.72 | 0.67 | 0.79 | 1.05E-13 | 4.15E-11 | A02BC Proton pump inhibitors |
| 1yrs | F | A02BC | 0.77 | 0.71 | 0.83 | 2.01E-11 | 4.16E-09 | A02BC Proton pump inhibitors |
| 2yrs | F | A02BC | 0.80 | 0.74 | 0.86 | 2.94E-09 | 4.17E-07 | A02BC Proton pump inhibitors |
| 0.5yr | G | A02BC | 0.70 | 0.61 | 0.81 | 1.06E-06 | 4.18E-05 | A02BC Proton pump inhibitors |
| 1yrs | G | A02BC | 0.66 | 0.58 | 0.76 | 1.56E-09 | 1.61E-07 | A02BC Proton pump inhibitors |
| 2yrs | G | A02BC | 0.68 | 0.59 | 0.77 | 1.81E-09 | 1.92E-07 | A02BC Proton pump inhibitors |
| 2yrs | F | A03FA | 0.51 | 0.37 | 0.70 | 3.67E-05 | 1.95E-03 | A03FA Propulsives |
| 1yrs | F | A09AA | 0.24 | 0.09 | 0.64 | 4.19E-03 | 6.94E-02 | A09AA Enzyme preparations |
| 2yrs | F | A09AA | 0.23 | 0.09 | 0.60 | 2.81E-03 | 5.70E-02 | A09AA Enzyme preparations |
| 0.5yr | F | A12AX | 0.80 | 0.69 | 0.93 | 2.74E-03 | 6.28E-02 | A12AX Calcium, combinations with vitamin D and/or other drugs |
| 1yrs | F | A12AX | 0.83 | 0.72 | 0.94 | 4.36E-03 | 6.94E-02 | A12AX Calcium, combinations with vitamin D and/or other drugs |
| 1yrs | F | B03AA | 0.74 | 0.60 | 0.91 | 4.00E-03 | 6.94E-02 | B03AA Iron bivalent, oral preparations |
| 0.5yr | F | G03CA | 0.63 | 0.52 | 0.76 | 3.03E-06 | 2.39E-04 | G03CA Natural and semisynthetic estrogens, plain |
| 1yrs | F | G03CA | 0.67 | 0.58 | 0.78 | 4.08E-07 | 4.22E-05 | G03CA Natural and semisynthetic estrogens, plain |
| 2yrs | F | G03CA | 0.70 | 0.61 | 0.80 | 1.89E-07 | 1.61E-05 | G03CA Natural and semisynthetic estrogens, plain |
| 2yrs | G | G03CA | 0.66 | 0.51 | 0.86 | 2.43E-03 | 6.08E-02 | G03CA Natural and semisynthetic estrogens, plain |
| 0.5yr | F | G04CB | 0.63 | 0.46 | 0.85 | 3.02E-03 | 6.28E-02 | G04CB Testosterone-5-alpha reductase inhibitors |
| 1yrs | C | H03AA | 0.62 | 0.48 | 0.79 | 1.77E-04 | 1.43E-03 | H03AA Thyroid hormones |
| 2yrs | C | H03AA | 0.62 | 0.48 | 0.79 | 1.51E-04 | 1.52E-03 | H03AA Thyroid hormones |
| 1yrs | E | H03AA | 0.80 | 0.71 | 0.92 | 9.47E-04 | 8.05E-03 | H03AA Thyroid hormones |
| 2yrs | E | H03AA | 0.80 | 0.70 | 0.91 | 5.94E-04 | 5.02E-03 | H03AA Thyroid hormones |
| 0.5yr | F | H03AA | 0.80 | 0.69 | 0.92 | 2.24E-03 | 5.53E-02 | H03AA Thyroid hormones |
| 1yrs | F | H03AA | 0.81 | 0.71 | 0.93 | 2.51E-03 | 5.20E-02 | H03AA Thyroid hormones |
| 2yrs | F | H03AA | 0.81 | 0.71 | 0.93 | 2.57E-03 | 5.47E-02 | H03AA Thyroid hormones |
| 0.5yr | G | H03AA | 0.66 | 0.51 | 0.86 | 2.10E-03 | 3.60E-02 | H03AA Thyroid hormones |
| 1yrs | G | H03AA | 0.64 | 0.49 | 0.82 | 5.53E-04 | 1.34E-02 | H03AA Thyroid hormones |
| 2yrs | G | H03AA | 0.64 | 0.50 | 0.83 | 6.06E-04 | 1.72E-02 | H03AA Thyroid hormones |
| 1yrs | F | J01EA | 0.69 | 0.53 | 0.90 | 6.09E-03 | 9.00E-02 | J01EA Trimethoprim and derivatives |
| 1yrs | F | J01MA | 0.49 | 0.34 | 0.72 | 2.40E-04 | 1.04E-02 | J01MA Fluoroquinolones |
| 2yrs | F | J01MA | 0.59 | 0.46 | 0.76 | 5.39E-05 | 2.55E-03 | J01MA Fluoroquinolones |
| 0.5yr | F | L02AE | 0.29 | 0.14 | 0.60 | 9.85E-04 | 3.24E-02 | L02AE Gonadotropin releasing hormone analogues |
| 1yrs | F | L02AE | 0.41 | 0.23 | 0.72 | 2.02E-03 | 4.58E-02 | L02AE Gonadotropin releasing hormone analogues |
| 2yrs | F | L02AE | 0.42 | 0.25 | 0.70 | 9.73E-04 | 2.44E-02 | L02AE Gonadotropin releasing hormone analogues |
| 0.5yr | F | M01AE | 0.68 | 0.56 | 0.82 | 4.61E-05 | 2.28E-03 | M01AE Propionic acid derivatives |
| 1yrs | F | M01AE | 0.79 | 0.70 | 0.91 | 6.65E-04 | 2.24E-02 | M01AE Propionic acid derivatives |
| 0.5yr | F | N02AX | 0.56 | 0.41 | 0.76 | 1.88E-04 | 7.43E-03 | N02AX Other opioids |
| 1yrs | F | N02AX | 0.63 | 0.49 | 0.80 | 1.64E-04 | 8.49E-03 | N02AX Other opioids |
| 2yrs | F | N02AX | 0.68 | 0.56 | 0.83 | 1.14E-04 | 3.74E-03 | N02AX Other opioids |
| 0.5yr | F | N03AX | 0.68 | 0.58 | 0.81 | 1.72E-05 | 9.71E-04 | N03AX Other antiepileptics |
| 1yrs | F | N03AX | 0.70 | 0.60 | 0.82 | 1.00E-05 | 6.90E-04 | N03AX Other antiepileptics |
| 2yrs | F | N03AX | 0.73 | 0.64 | 0.84 | 7.15E-06 | 5.08E-04 | N03AX Other antiepileptics |
| 0.5yr | F | N06AA | 0.77 | 0.65 | 0.92 | 3.99E-03 | 7.51E-02 | N06AA Non-selective monoamine reuptake inhibitors |
| 1yrs | F | N06AA | 0.79 | 0.68 | 0.92 | 1.98E-03 | 4.58E-02 | N06AA Non-selective monoamine reuptake inhibitors |
| 2yrs | F | N06AA | 0.79 | 0.70 | 0.90 | 2.67E-04 | 8.12E-03 | N06AA Non-selective monoamine reuptake inhibitors |
| 1yrs | A | R03BA | 0.48 | 0.31 | 0.73 | 7.44E-04 | 1.35E-02 | R03BA Glucocorticoids |
| 2yrs | A | R03BA | 0.55 | 0.38 | 0.81 | 2.44E-03 | 5.64E-02 | R03BA Glucocorticoids |
| 0.5yr | F | R05DA | 0.69 | 0.55 | 0.87 | 1.46E-03 | 4.12E-02 | R05DA Opium alkaloids and derivatives |
| 1yrs | F | R05DA | 0.74 | 0.62 | 0.88 | 5.47E-04 | 2.06E-02 | R05DA Opium alkaloids and derivatives |
| 2yrs | F | R05DA | 0.80 | 0.70 | 0.91 | 7.02E-04 | 1.99E-02 | R05DA Opium alkaloids and derivatives |

Only results with FDR<0.1 are shown. Ophthalmological and dermatological agents are not listed in the above table.

#### Drugs ranked by consistency of protective associations

We also ranked the drugs in term of their *consistency* of protective associations. Briefly, drugs were ranked by their frequency of being nominally significant (p<0.05) across the 5 time-windows and 8 models (Table 6). This serves as a rough reference to prioritize drugs with potential protective effects. For some drugs, the results may not be significant after FDR correction. Nevertheless, if a drug showed consistent associations (at least nominally) across multiple models or time-frames, it may also be worthy of further investigations.

**Table 6.** Drugs showing consistent protective associations across 4 time-windows and 8 models (ranked by the frequency of being nominally significant i.e. p<0.05)

|  | ATC code | Drug name | Freq |
| --- | --- | --- | --- |
| 1 | C09AA | ACE inhibitors, plain | 21 |
| 2 | J07BB | Influenza vaccines | 20 |
| 3 | C10AA | HMG CoA reductase inhibitors | 19 |
| 4 | H03AA | Thyroid hormones | 17 |
| 5 | C09CA | Angiotensin II receptor blockers, plain | 15 |
| 6 | G04CB | Testosterone-5-alpha reductase inhibitors | 12 |
| 7 | A02BC | Proton pump inhibitors | 11 |
| 8 | C08CA | Dihydropyridine derivatives | 11 |
| 9 | R03BA | Glucocorticoids | 9 |
| 10 | C07AB | Beta blocking agents, selective | 8 |
| 11 | A10BA | Biguanides | 7 |
| 12 | B01AC | Platelet aggregation inhibitors excl. heparin | 7 |
| 13 | G03CA | Natural and semisynthetic estrogens, plain | 7 |
| 14 | J07CA | Bacterial and viral vaccines, combined | 7 |
| 15 | A03AA | Synthetic anticholinergics, esters with tertiary amino group | 6 |

#### Drug associated with increased odds of risk/severity of infection

Among the drugs with harmful associations, the more frequently top-listed ones include laxatives, opioids (N02AA), benzodiazepines, tetracycline, penicillins, other antipsychotics (N05AX) and anti-dementia drugs (N06DA/DX). The full results are presented in Table S6-10 and a summary is also provided in Table S5.

### Analysis restricted to subjects with complete covariate data, and models with/without IPW

As a sensitivity analysis, for the above analysis with imputed covariates, we also repeated models A to H *without* IPW of Pr(tested). In addition, we have also repeated the analyses limiting to subjects with complete covariate data, with or without the IPW approach. In general, we observed similar drugs with significant results and the top-ranked protective or harmful drugs were similar to the above. Comparing results with and without IPW, the list of significant drugs remained similar although the OR estimates and SE were adjusted. The full results are presented in Table S11-12 (complete covariate data with and without IPW) and S13 (imputed covariates without IPW).

### Subgroup analysis

A summary is presented in Table 7. The proportion of subjects falling in each subgroup is presented in Table S14 while full results are presented in Tables S15. We performed a statistical test to compare the log(OR) across the two subgroups with and without the risk factor; drugs with protective effect in one subgroup but significantly different OR in the other subgroup are listed in Table 7. For example, the protective effects of pneumococcal and flu vaccines were significantly weaker in obese (BMI>30) subjects under model F. With regards to age, several drugs, PPI and ACEI showed larger protective effects in those with age>70 under models F and E respectively. Statins, ACEIs and PPI showed stronger protective associations in HT under models C, E and F respectively. Regarding ethnicity as a subgroup, a number of drugs, including several vaccines, appeared to have stronger protective effects in the white compared to non-white subjects. However, only <10% of the UKBB subjects included here were non-white, and the non-white subgroup was heterogeneous and composed of several different ethnicities. We did not observe clear evidence of sex-specific effects in this analysis.

**Table 7.** Summary of subgroup analysis, showing drugs having significant association in one subgroup but significantly different OR in the other subgroup (FDR<0.2)

| Subgp | Windows | Model | OR_Y | OR_N | sig_Y | sig_N | z_OR_cmp | p_OR_cmp | p.adjust_OR_cmp | Name |
| --- | --- | --- | --- | --- | --- | --- | --- | --- | --- | --- |
| AGE>70 | p5yr | F | 0.81 | 0.99 | 1 | 0 | -2.65 | 8.15E-03 | 1.47E-01 | A02BC Proton pump inhibitors |
| AGE>70 | p1yr | E | 1.19 | 0.49 | 0 | 1 | 2.11 | 3.47E-02 | 1.56E-01 | A10AE Insulins and analogues for injection, long-acting |
| AGE>70 | p1yr | E | 0.81 | 1.04 | 1 | 0 | -2.38 | 1.72E-02 | 1.55E-01 | C09AA ACE inhibitors, plain |
| Asthma | p5yr | E | 0.60 | 0.86 | 1 | 1 | -2.66 | 7.76E-03 | 1.40E-01 | J07BB Influenza vaccines |
| Asthma | p10y | E | 0.61 | 0.87 | 1 | 1 | -2.95 | 3.22E-03 | 5.80E-02 | J07BB Influenza vaccines |
| BMI>30 | p1yr | F | 1.04 | 0.31 | 0 | 1 | 2.42 | 1.56E-02 | 1.40E-01 | J07AL Pneumococcal vaccines |
| BMI>30 | p1yr | F | 0.76 | 0.54 | 1 | 1 | 2.52 | 1.17E-02 | 1.40E-01 | J07BB Influenza vaccines |
| BMI>30 | p2yr | F | 0.79 | 0.56 | 1 | 1 | 2.75 | 6.01E-03 | 1.08E-01 | J07BB Influenza vaccines |
| BMI>30 | p6m | F | 0.92 | 0.16 | 0 | 1 | 2.68 | 7.40E-03 | 1.26E-01 | R03BA Glucocorticoids |
| CAD | p5yr | H | 0.36 | 1.32 | 1 | 0 | -2.39 | 1.71E-02 | 1.53E-01 | C08CA Dihydropyridine derivatives |
| CAD | p5yr | H | 1.72 | 0.18 | 0 | 1 | 2.42 | 1.53E-02 | 1.53E-01 | G04CB Testosterone-5-alpha reductase inhibitors |
| CAD | p5yr | C | 1.92 | 0.56 | 0 | 1 | 2.38 | 1.72E-02 | 1.55E-01 | J07AL Pneumococcal vaccines |
| CAD | p5yr | F | 1.55 | 0.56 | 0 | 1 | 2.55 | 1.07E-02 | 1.92E-01 | J07AL Pneumococcal vaccines |
| Depression | p1yr | B | 0.07 | 0.73 | 1 | 0 | -2.60 | 9.36E-03 | 1.50E-01 | C10AA HMG CoA reductase inhibitors |
| HT | p2yr | F | 0.75 | 0.93 | 1 | 0 | -2.69 | 7.20E-03 | 1.30E-01 | A02BC Proton pump inhibitors |
| HT | p5yr | F | 0.76 | 1.00 | 1 | 0 | -3.36 | 7.92E-04 | 1.43E-02 | A02BC Proton pump inhibitors |
| HT | p1yr | E | 0.86 | 1.07 | 1 | 0 | -2.11 | 3.49E-02 | 1.26E-01 | C09AA ACE inhibitors, plain |
| HT | p1yr | C | 0.71 | 1.02 | 1 | 0 | -2.58 | 9.90E-03 | 8.91E-02 | C10AA HMG CoA reductase inhibitors |

**Table 8.**
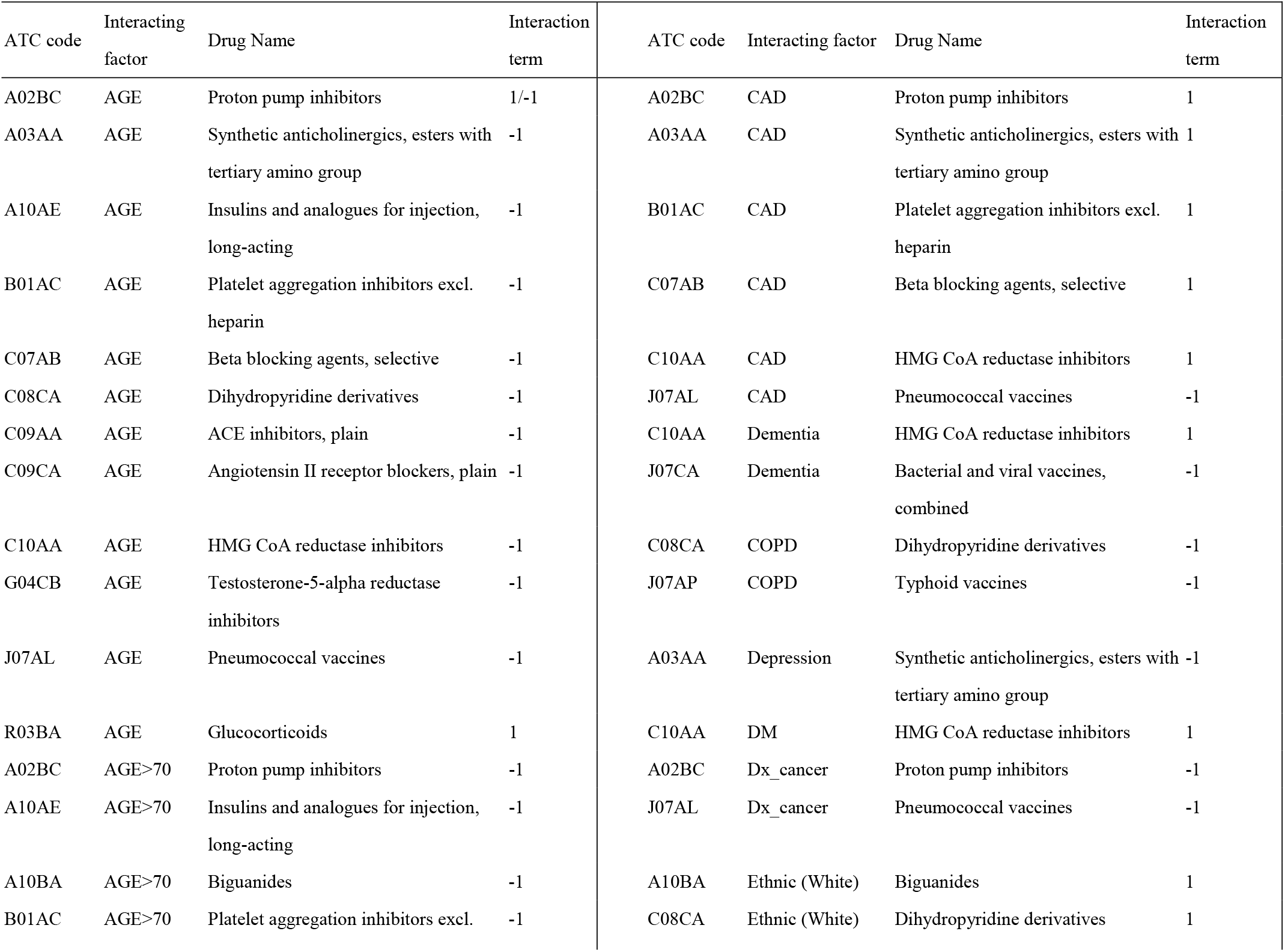
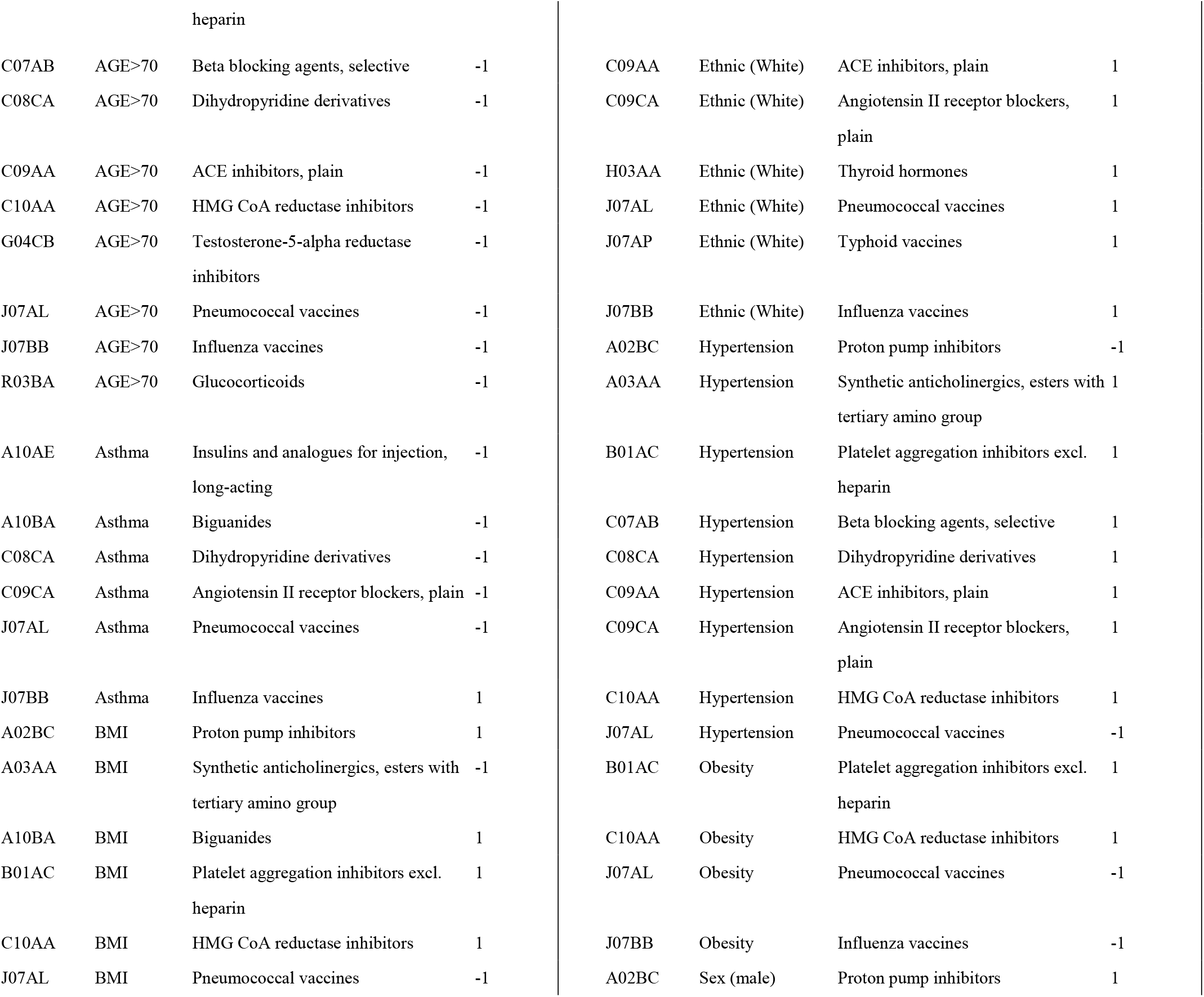
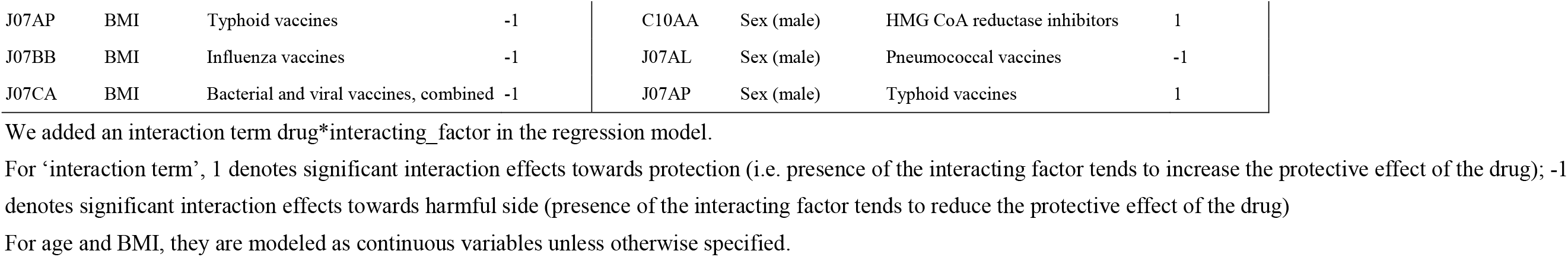
Summary of interaction analysis, showing pairs of variables with significant interactions (FDR<0.2)

### Interaction analysis

A summary of results (results with FDR<0.2) is presented in Table S8 while a fuller version is given in Table S16. Full results are given in Tables S17. More significant results at FDR<0.2) are observed, presumably due to higher power of the interaction-term approach. For example, we found that most vaccines showing protective effects, including influenza and pneumococcal vaccines, interacted with BMI and obesity significantly. Higher BMI was associated with *reduced* protective effects, in line with evidence from subgroup analysis. On the other hand, statins, biguanides (metformin) and anti-platelet drugs showed positive interactions with BMI. For CAD, significant interaction was observed with several CM drugs including beta-blockers (non-selective), antiplatelet drugs and statins, suggesting larger protective effects for such drugs in CAD patients. In a similar vein, most CM medications showed interaction with HT indicating more prominent protective associations in HT patients. Considering age as an interacting variable, interaction was observed with a large number of drugs, most suggesting weaker protection as age increases.

From the perspective of specific medications, for example, statins interact with multiple risk factors and demonstrate larger protective effects with CAD, obesity, DM, CAD, HT, dementia and in males. However, its effect tends to be weaker with increasing age. Interaction analysis with flu vaccines showed that its effect may be weaker in the obese and with increasing age, but was stronger in the white population and asthmatic subgroup. ACEI and ARB showed stronger protective effects in the white and HT patients, but weaker effects with advanced age.

### Controlling for other medications

We primarily focused on protective drugs as the number of drugs with significant negative effects is large and is hard to control for all. Overall speaking, most drugs with protective effects remain significant (at least for a subset of models) despite controlling for other medications (Table S19). However, biguanides (A10BA), CCB (C08CA) and platelet aggregation inhibitors excluding heparin (B01AC) showed a consistent trend of non-significant association with the outcome when other protective drugs were controlled for. The findings are similar when controlling for top 10/20 drugs or all protective drugs having FDR<0.05/0.1.

## Discussion

In this work, we have performed a thorough and rigorous analysis on the effect of drugs and vaccines on COVID-19 susceptibility and severity. We uncovered a number of drugs with potentially protective or harmful effects.

As an observational study, different kinds of bias such as confounding and selection bias may affect the results. We have performed analysis on infected subjects (models A and B), the whole population (models C, D, E) and the tested population (models F, G, H) to obtain a more comprehensive picture of drug effects under different settings, and to avoid limitations (e.g. selection/collider bias) of some designs.

We note that sometimes the different models may yield different results. One main observation is that analysis on the tested population appears to result in more findings of drugs with protective effects. We also observed that some drugs in model F (infected vs tested negative) may show different effects under model E (infected vs general population). Several reasons may explain this finding. First and foremost, confounding by indication is inevitable and may play a more important role when analyzing general population samples. It is possible that apparent harmful effects of drugs are due to the diseases/conditions that the prescription is related to, or to poorer health in general.

Based on an machine learning model to predict testing probability (see Figure S1), we observed that people who are older, having more comorbidities and taking more medications, suffering from cardiovascular conditions etc. were more likely to be tested. Compared to the general population, the tested group may represent a more ‘homogeneous’ population, enriched for people with poorer health and more comorbidities in general. Therefore a proportion of confounders, which overlap with factors associated with higher probability of being tested, are essentially controlled for by stratification if we only study the tested subjects. On the other hand, in the general population, as there is a higher proportion of healthy subjects, the effect of confounding by indication may be stronger. Another possibility is collider bias due to conditioning on a subgroup of subjects. For example, a drug may be associated with certain conditions which in turn are associated with higher chance of being tested; on the other hand those who have more severe symptoms or complications are more likely to be tested. Conditioning on testing may result in spurious associations between the drug and severity of infection. However, we have tried to minimize this type of bias by the IPW approach, and we did not observe substantial difference in results with or without IPW correction for most drugs. However, we note that even with adjustment by IPW, there is still chance for residual selection or collider bias. For example, some factors associated with Pr(tested) may not be captured in the prediction model. A third possibility to consider is that a drug may truly produce different effects in different subgroups, due to effect modification by other factors or diseases. For instance, a recent study reported that the protective effect of statins is more marked in patients with diabetes ^22^. The fact that risk factor associations may differ between a whole-population or tested-population based study has also been noted previously, for example by ^17^.

### Highlights of relevant drugs

Below we highlight drugs that are tentatively associated with altered risk or severity of infection. We will preferentially consider drugs that showed significant associations (with FDR<0.1) across multiple models and time-windows, those with stronger statistical significance, and those with protective effects as confounding by indication is much less likely.

#### Drugs for cardiometabolic disorders with protective effects

Interestingly, many drugs with potential protective effects are indicated for cardiometabolic (CM) disorders. Cardiometabolic risk factors, such as obesity, hypertension, DM and CAD, have consistently been shown to be related to risk and severity of infection; as such, it is biologically plausible that drugs for treating CM disorders may be beneficial.

Among all drugs, the strongest and most consistent protective association was observed for statins. The beneficial effects of statins were supported by several previous studies. For example, a recent meta-analysis of four retrospective studies of COVID-19 patients ^23^ showed a significantly decreased hazard of severity or mortality of infection (pooled HR= 0.70) when comparing statin users against non-users. Another retrospective study by Tan et al. ^24^ also reported lower risk of ICU admission among statin users in infected patients. Yet another work showed that statins may be effective in reducing in-hospital mortality among diabetic patients ^22^. Potential mechanisms for the protective actions of statins have been discussed elsewhere ^25-27^. It has been postulated that besides reducing CVD risks, statins may reduce risk/severity of infection by inhibiting inflammation and excessive immune response, producing direct antiviral effects, improving endothelial function and exerting an antithrombotic effect, among other actions^25-27^.

Another group of drugs worth highlighting is ACEI and ARB. There have been intense discussions on whether ACEI/ARB may affect risk or severity of infection from early on, as ACE2 is a receptor for SARS-CoV-2. Nevertheless, a recent study showed that ACE2 is localized in respiratory cilia, and the use of ARB/ACEI does not change its expression. ^28^. Recent systemic reviews and meta-analysis (for example see ^29^ with continuous updates) of observational studies do not support an association between ACEI/ARB prior use and severity of infection. However, several studies ^28,30-36^ reported protective effects of ACEI/ARB on severity or mortality of disease. Here we observed highly consistent association of prior use of ACEI/ARB on reduced risks of severe/fatal infection (models A, C, G), and overall infection risk in the population (model E).

For several other kinds of CM drugs, the associations are not as strong but may still be worthy of further studies. Biguanides (mainly metformin) are observed to be protective for severe COVID-19 infection, both among the infected and at a population level. For example, in a meta-analysis on four observational studies of hospitalized patients mostly with type 2 DM, the use of metformin was associated with a lower risk of mortality (OR = 0.75, 95% CI = 0.67-0.85) ^37^. A number of mechanisms have been proposed ^37,38^. For example, besides improving glycemic control and weight reduction, metformin may lead to AMPK activation which potentially reduces viral entry by phosphorylation of ACE2 receptor. It may also lead to mTOR pathway inhibition and prevents hyperactivation of the immune system ^37^.

Other drugs of interest may include beta-blockers and calcium channel blockers (C08CA, dihydropyridine derivatives). It was suggested that beta-blockers may be useful in preventing hyperinflammation and hence beneficial for COVID-19 ^39^. For calcium channel blockers (CCBs), a study using cell culture suggested that CCBs, especially amlodipine and nifedipine, were useful in blocking viral entry and infection in epithelial lung cells. ^40^. In another retrospective study ^41^, both beta-blockers and CCBs were associated with lower mortality. Another relevant study in the UK ^42^ utilized data from the UK Clinical Practice Research Datalink (CPRD) and found that ACEI/ARB, CCBs and thiazide diuretics were all associated lower odds of diagnosis, while beta-blockers do not show any association after adjusting for consultation frequency. None of the above drugs were associated with mortality in that study ^42^.

#### Vaccines

There has been intense interest in whether vaccines indicated for other diseases may protect against COVID-19. Here we observed that a number of vaccines (ATC code J07) showed protection against infection or severe infection. For example, pneumococcal vaccines were protective against infection in the population and tested subjects, and risk of severe infection (model G). Significant protective associations were also observed for tetanus and typhoid vaccines at a time horizon of 10 years (the power to detect associations is likely stronger over longer periods due to larger number of people having received the vaccine; it does not exclude the possibility that the vaccines may have effects over shorter time windows). We also observed associations with J07CA category, which contains various bacterial and viral vaccines (see https://www.whocc.no/atc_ddd_index/?code=J07CA).

For influenza vaccines, we observed highly consistent protective associations. It has been proposed that ‘trained innate immunity’, which may involve epigenetic reprogramming of innate immune cells, may enable a vaccine to protect against other diseases ^43,44^. Interestingly, two studies in Italy reported that higher coverage rate of flu vaccine was associated with lower rate of infection, hospitalization and mortality from COVID-19. Another larger-scale study based on electronic records of 137,037 subjects who have received viral PCR tests showed that a number of vaccines (given in the past 1, 2 or 5 years) were associated with lower risks of infection ^45^. These included flu and pneumococcal vaccines also implicated in the present study. Yet another recent study in the Netherlands ^46^ also showed a reduce risk [RR = 0.61 (95% CI, 0.46 - 0.82)] of infection among recipients of flu vaccine, and this effect size was similar to what we observed from this study. In vitro studies by the same authors showed that the vaccine is able to induce a trained immunity response, including an increase of cytokine responses after stimulation of immune cells with SARS-CoV-2.

We note that this is an observational study and residual confounding may be present. For example, it is possible that people receiving flu vaccines are more health-conscious and observe preventive measures better. However, we observed waning protective effects over time, which makes sense biologically but may not be entirely explained by the above confounder as the level of health-consciousness tend not to change over a few years’ time. Also, the vaccine appears to have stronger effect sizes if fatal infection is considered as the outcome (although the confidence interval is large), which cannot be easily explained by health-consciousness. On the other hand, as flu vaccines are more likely to be received by the elderly and those with chronic illnesses, residual confounding of these factors tend to push the effects towards the harmful side.

Taken together, we believe that the protective effects of vaccines may not be easily explained away by other confounders. Further experimental and clinical studies are warranted to investigate the non-specific effects of flu and other vaccines, especially since COVID-19 vaccines may not be easily available to many people in a short time-frame.

#### Other potential protective drugs

We briefly highlight a few other drugs with potential protective effects. Estrogens (G03CA) were among the drugs showing protective associations. As many studies reported higher risks of severe disease in men than in women, it has been hypothesized that estrogen may play a part in the sex-discordant outcomes, for example via its effects on immune response to infections ^47-49^. Thyroid hormones (TH) were also among the top-ranked drugs. It was postulated that TH may ameliorate tissue injury due to hypoxia by suppression of p38 MAPK ^50^. Clinical trials on TH are ongoing^50,51^ and our findings support a protective role of TH in COVID-19. Another drug category of note is proton pump inhibitors (PPI). Several studies have suggested harmful effects of PPI on disease severity, which may be related to reduced gastric acid production with subsequent bacterial overgrowth ^52-54^. However, an in-vitro screening study revealed that PPIs may serve as a potent inhibitor of SARS-CoV-2 replication ^55^. The difference in findings between the current study and previous works may be due to heterogeneity in study samples and designs, differences in the outcome studied (e.g. hospitalization vs ICU admission used in some other studies; infection risk vs severity of disease etc.) and variations in the covariates being adjusted. Residual confounding, such as by other comorbidities and drugs given, may also affect the results. Interestingly, we observed that effects of PPI may be stronger in certain subgroups (e.g. older age, HT), which may also account for the discrepancy in results in different studies.

Several other top-ranked drug categories in Table 6 may be worth discussing. Testosterone-5-alpha reductase inhibitors (5ARis) were recently shown in a small RCT to reduce the time to remission ^56^. Two earlier observational studies also reported lower risk of ICU admission and frequency of symptoms ^57,58^. 5ARis blocks the conversion of testosterone to its more potent form, dihyrotestosterone. Of note, one of the key receptors for the SAR-CoV-2 virus is TMPRSS2 ^59^, and the only known promoter of the genes is an androgen response element in the promoter region ^60^. Another drug category is platelet aggregation inhibitors (B01AC). It has been reported that COVID-19 is associated with higher risk of thrombotic events including deep vein thrombosis and pulmonary embolism ^61^. Anti-thrombotic therapies have been hypothesized to reduce thrombo-inflammatory processes as a result of endothelial dysfunction related to viral infection ^62^. An observational study reported that aspirin is associated with reduced risk of mechanical ventilation and mortality in hospitalized patients ^63^, however RCTs are lacking.

For some of the protective drugs highlighted above, we note that their significance weakened (or became non-significant) when controlling for other medications. However, we expect multi-collinearity among the drug variables, as cardiometabolic disorders are highly comorbid and one patient often takes multiple medications. Multi-collinearity may render interpretation of individual predictors difficult due to unstable coefficient estimates. ^64^

In our secondary analyses we also considered **subgroup and interaction effects**. While this is a more exploratory analysis and further replications are required, it shed light on how the effects of drugs/vaccines may differ in people with different clinical background. For instance, we observed a consistent trend that the protective associations of flu and pneumococcal vaccines were weaker in obese individuals. As an example, comparing those who received flu vaccine in the past season (2019-2020) against those who did not, the estimated OR for infection was 0.76 in the obese group and 0.54 in the non-obese group (model F). It has been observed before that obese individuals responded less well to flu and other vaccines due to impaired immunological responses^65,66^. As another example, statins were observed to have more prominent protective effects in those with cardiometabolic abnormalities, such as DM, HT, CAD and obesity. This is also supported by a recent study ^22^ which showed mortality reduction in statin users in diabetic patients only.

#### Drugs with potentially harmful effects

We noted a number of drugs with potentially harmful effects, but we caution that residual confounding, such as confounding by indication, other comorbidities and general poor health, may lead to bias towards an increased odds of infection or severe disease.

For example, people who have poorer health in general may visit their GPs more often and be prescribed drugs (e.g. laxatives, antibiotics, painkillers), which may lead to confounding. Nevertheless, it is possible that some of the top-ranked drugs may indeed increase the risk/severity of infection. For instance, it is slightly unexpected that laxatives were highly significant across multiple models and time windows. Of note, it has recently been postulated that dysregulation of gut microbiome may be associated with susceptibility or resilience to infection ^67,68^, and laxatives represent a main category of drugs that affects the gut microbiome ^69^. Interestingly, several associations involve psychiatric medications such as benzodiazepines, antipsychotics and anti-dementia drugs. The association may be due to underlying neuropsychiatric conditions (e.g. anxiety, psychosis, dementia etc.), or the effect of the drugs, or a combination of both. Some of above drugs overlapped with those revealed in a recent study using primary care data in Scotland. In a univariate analysis restricted to non-residents in care homes and those without major conditions, laxatives, anxiolytics, penicillins and opioid analgesics were significantly associated with ICU admission or mortality from COVID-19 when compared to population controls ^70^. These drugs were also top-listed as those with harmful effects in our study.

### Strengths and limitation

This study has a number of strengths. First and foremost, the study was performed on a large cohort of subjects with a sample size close to half a million. The sample was not limited to one or a few medical centers and covered the entire UK population, although this is not an entirely random sample and participation bias still exists ^16^. The large and well-characterized sample also enables analysis of infected, tested as well the whole population. We have studies all level-4 ATC drug categories, allowing an unbiased and systematic analysis on the association of different drugs with COVID-19 risks or outcomes. This avoids the risk of publication bias, especially negative results to be unreported. Drugs showing null associations are still be of important public health interest, as this may suggest that patients on such medications may not need to change their regimen in view of the pandemic. Medication history was retrieved from GP records, which minimize recall bias and errors from self-reporting. Another strength is that we performed a variety of statistical analysis to reduce bias, including control for potential confounders, multiple imputation, IPW to reduce effects of testing bias, and study of different time windows and multiple models. Some of our findings were corroborated by previous studies; however, many previous clinical studies were limited to hospitalized or infected individuals, which cannot study the effect of drugs on susceptibility to infection. Selection on hospitalized/infected subjects may be prone to selection/collider bias as discussed elsewhere ^16^, therefore we included multiple models with infected, tested as well the whole population as samples, which aims to reduce bias and limitations due to specific study designs.

There are also various limitations, some of which are mentioned above. First and foremost, this is an observational study based on a retrospective cohort of UKBB. As this is not a randomized controlled trial (RCT), confounding is inevitable, especially confounding by indication. Although we have controlled for main confounders in the regression model, residual confounding is still likely. Since confounding by indication will likely bias towards *increased* odds of infection or severe disease, null or protective associations may be more reliable. Confounding by the use of other types of drugs is also possible. Also, the UKBB cohort is not random and participants are in general healthier than the general population ^71^. The majority of participants are of European descent so the findings may not be generalizable to other ethnicities. Also, the subjects are mostly >50 years old and drug effects in younger individuals may be different.

Regarding drug history, it is worth noting that vaccination records are not complete as individuals may receive vaccination outside GP practices. Over-the-counter prescriptions were not counted, and it cannot be guaranteed that all drugs issued are dispensed by the pharmacy (https://biobank.ctsu.ox.ac.uk/crystal/crystal/docs/tppgp4covid19.pdf). However, if this misclassification is non-differential (unrelated to outcome), the bias will be towards the null. There is a relatively high missing rate of GP prescription records for deceased COVID-19 patients, which leads to reduced power to detect associations. While the UKBB cohort sample is large, we still have low power to detect associations for drugs that are uncommonly prescribed. Another limitation with the GP records is that only the issue date but no duration or dosage is available.

As for the outcome, hospitalization is a rough proxy for severity only. For models comparing to the general population, it is likely that a proportion of the population may be infected but were not tested. This tends to lead to bias on the conservative side (akin to the use of unscreened controls in genetic stuides ^72,73^), especially under model E. Patients with more severe symptoms are less likely to remain untested, so other models may be less affected by this bias. Finally, this study focuses on prior (or pre-diagnostic) use of drugs and their association with infection risk/severity, and does not directly address whether newly prescribed drugs to recently diagnosed patients will be useful or not.

## Conclusions

Here we observed that a number of drugs, including many for cardiometabolic disorders, may be associated with lower odds of infection/severity of COVID-19. Several existing vaccines, especially flu vaccines, may be beneficial against COVID-19 as well. Due to the observational nature of the study, confounding cannot be excluded, and other bias and limitations may be present. We understand that causal relationship between drugs and disease cannot be reliably concluded from this study alone, and shall regard the findings as more exploratory than confirmatory. Nevertheless, being one of the most comprehensive studies to date on drug associations, we believe the current work provided a valuable resource to prioritize relevant drugs for future meta-analyses, clinical trials or experimental studies.

## Data Availability

The UK Biobank data is available to registered researchers.

## Acknowledgements

This work was supported partially by the Lo Kwee Seong Biomedical Research Fund from The Chinese University of Hong Kong and an NSFC grant. We thank Prof. Pak Sham for support on data access. We thank Mr Carlos Chau for help with data presentation, Ms Liangying YIN for help with data cleaning and Dr Shitao RAO and Ms Jinghong QIU for help in manuscript preparation.

## Supplementary Information

All supplementary Tables and notes are available at https://drive.google.com/drive/folders/1_noITkBAsef_7Kb6bUd_RI_3VQK5jafH?usp=sharing

## References

1. Li, Q. et al. Early Transmission Dynamics in Wuhan, China, of Novel Coronavirus-Infected Pneumonia. N Engl J Med (2020).

2. Novel-Coronavirus-Pneumonia-Emergency-Response-Epidemiology-Team. [The epidemiological characteristics of an outbreak of 2019 novel coronavirus diseases (COVID-19) in China]. Zhonghua Liu Xing Bing Xue Za Zhi 41, 145–151 (2020).

3. Guan, W.-j. et al. Clinical Characteristics of Coronavirus Disease 2019 in China. New England Journal of Medicine (2020).

4. Kwok, S. et al. Obesity: A critical risk factor in the COVID-19 pandemic. Clinical Obesity 10, e12403 (2020).

5. Zhou, F. et al. Clinical course and risk factors for mortality of adult inpatients with COVID-19 in Wuhan, China: a retrospective cohort study. The Lancet 395, 1054–1062 (2020).

6. Maddaloni, E. et al. Cardiometabolic multimorbidity is associated with a worse Covid-19 prognosis than individual cardiometabolic risk factors: a multicentre retrospective study (CoViDiab II). Cardiovascular Diabetology 19, 164 (2020).

7. Gansevoort, R.T. & Hilbrands, L.B. CKD is a key risk factor for COVID-19 mortality. Nature Reviews Nephrology 16, 705–706 (2020).

8. Zhou, Y. et al. Comorbidities and the risk of severe or fatal outcomes associated with coronavirus disease 2019: A systematic review and meta-analysis. International Journal of Infectious Diseases 99, 47–56 (2020).

9. Marigorta, U.M., Rodríguez, J.A., Gibson, G. & Navarro, A. Replicability and Prediction: Lessons and Challenges from GWAS. Trends in genetics : TIG 34, 504–517 (2018).

10. Sudlow, C. et al. UK Biobank: An Open Access Resource for Identifying the Causes of a Wide Range of Complex Diseases of Middle and Old Age. PLOS Medicine 12, e1001779 (2015).

11. Antia, A. et al. Heterogeneity and longevity of antibody memory to viruses and vaccines. PLOS Biology 16, e2006601 (2018).

12. Benjamini, Y. & Hochberg, Y. Controlling the False Discovery Rate: A Practical and Powerful Approach to Multiple Testing. Journal of the Royal Statistical Society: Series B (Methodological) 57, 289–300 (1995).

13. Waljee, A.K. et al. Comparison of imputation methods for missing laboratory data in medicine. BMJ Open 3, e002847 (2013).

14. Wright, M.N. & Ziegler, A. ranger: A Fast Implementation of Random Forests for High Dimensional Data in C++ and R. Journal of Statistical Software; Vol 1, Issue 1 (2017) (2017).

15. Rubin, D.B. Multiple imputation for nonresponse in surveys, (John Wiley & Sons, 2004).

16. Griffith, G.J. et al. Collider bias undermines our understanding of COVID-19 disease risk and severity. Nature Communications 11, 5749 (2020).

17. Yates, T. et al. Framework to aid analysis and interpretation of ongoing COVID-19 research [version 1; peer review: 1 approved with reservations]. Wellcome Open Research 5(2020).

18. Wong, K.C.Y. & So, H.-C. Uncovering clinical risk factors and prediction of severe COVID-19: A machine learning approach based on UK Biobank data. medRxiv, 2020.09.18.20197319 (2020).

19. Kull, M., Silva Filho, T. & Flach, P. Beta calibration: a well-founded and easily implemented improvement on logistic calibration for binary classifiers. in Artificial Intelligence and Statistics 623–631 (2017).

20. Xu, S. et al. Use of stabilized inverse propensity scores as weights to directly estimate relative risk and its confidence intervals. Value Health 13, 273–7 (2010).

21. Buckley, J.P., Doherty, B.T., Keil, A.P. & Engel, S.M. Statistical Approaches for Estimating Sex-Specific Effects in Endocrine Disruptors Research. Environmental Health Perspectives 125, 067013 (2017).

22. Saeed, O. et al. Statin Use and In-Hospital Mortality in Diabetics with COVID-19. Journal of the American Heart Association 0, e018475 (2020).

23. Kow, C.S. & Hasan, S.S. Meta-analysis of Effect of Statins in Patients with COVID-19. American Journal of Cardiology 134, 153–155 (2020).

24. Tan, W.Y.T., Young, B.E., Lye, D.C., Chew, D.E.K. & Dalan, R. Statin use is associated with lower disease severity in COVID-19 infection. Scientific Reports 10, 17458 (2020).

25. Ganjali, S. et al. Commentary: Statins, COVID-19, and coronary artery disease: killing two birds with one stone. Metabolism: clinical and experimental 113, 154375–154375 (2020).

26. Minz, M.M., Bansal, M. & Kasliwal, R.R. Statins and SARS-CoV-2 disease: Current concepts and possible benefits. Diabetes & metabolic syndrome 14, 2063–2067 (2020).

27. Lee, K.C.H., Sewa, D.W. & Phua, G.C. Potential role of statins in COVID-19. International journal of infectious diseases : IJID : official publication of the International Society for Infectious Diseases 96, 615–617 (2020).

28. Lee, I.T. et al. ACE2 localizes to the respiratory cilia and is not increased by ACE inhibitors or ARBs. Nature Communications 11, 5453 (2020).

29. Mackey, K., Kansagara, D. & Vela, K. Update Alert 4: Risks and Impact of Angiotensin-Converting Enzyme Inhibitors or Angiotensin-Receptor Blockers on SARS-CoV-2 Infection in Adults. Annals of internal medicine 173, W147–W148 (2020).

30. Barochiner, J. & Martínez, R. Use of inhibitors of the renin-angiotensin system in hypertensive patients and COVID-19 severity: A systematic review and meta-analysis. J Clin Pharm Ther (2020).

31. Hippisley-Cox, J. et al. Risk of severe COVID-19 disease with ACE inhibitors and angiotensin receptor blockers: cohort study including 8.3 million people. Heart 106, 1503–1511 (2020).

32. Megaly, M. & Glogoza, M. Renin-angiotensin system antagonists are associated with lower mortality in hypertensive patients with COVID-19. Scott Med J 65, 123–126 (2020).

33. Pan, W. et al. Clinical Features of COVID-19 in Patients With Essential Hypertension and the Impacts of Renin-angiotensin-aldosterone System Inhibitors on the Prognosis of COVID-19 Patients. Hypertension 76, 732–741 (2020).

34. Bean, D. et al. ACE-inhibitors and Angiotensin-2 Receptor Blockers are not associated with severe SARS-COVID19 infection in a multi-site UK acute Hospital Trust. medRxiv, 2020.04.07.20056788 (2020).

35. Feng, Y. et al. COVID-19 with Different Severities: A Multicenter Study of Clinical Features. Am J Respir Crit Care Med 201, 1380–1388 (2020).

36. Zhang, P. et al. Association of Inpatient Use of Angiotensin-Converting Enzyme Inhibitors and Angiotensin II Receptor Blockers With Mortality Among Patients With Hypertension Hospitalized With COVID-19. Circ Res 126, 1671–1681 (2020).

37. Scheen, A.J. Metformin and COVID-19: From cellular mechanisms to reduced mortality. Diabetes & metabolism 46, 423–426 (2020).

38. Sharma, S., Ray, A. & Sadasivam, B. Metformin in COVID-19: A possible role beyond diabetes. Diabetes research and clinical practice 164, 108183–108183 (2020).

39. Barbieri, A. et al. Can Beta-2-Adrenergic Pathway Be a New Target to Combat SARS-CoV-2 Hyperinflammatory Syndrome?—Lessons Learned From Cancer. Frontiers in Immunology 11(2020).

40. Straus, M.R., Bidon, M., Tang, T., Whittaker, G.R. & Daniel, S. FDA approved calcium channel blockers inhibit SARS-CoV-2 infectivity in epithelial lung cells. bioRxiv, 2020.07.21.214577 (2020).

41. Chouchana, L. et al. Association of antihypertensive agents with the risk of in-hospital death in patients with Covid-19. medRxiv, 2020.11.23.20237362 (2020).

42. Rezel-Potts, E., Douiri, A., Chowienczyk, P.J. & Gulliford, M.C. Antihypertensive Medications and COVID-19 Diagnosis and Mortality: Population-based Case-Control Analysis in the United Kingdom. medRxiv, 2020.09.25.20201731 (2020).

43. Blok, B.A., Arts, R.J., van Crevel, R., Benn, C.S. & Netea, M.G. Trained innate immunity as underlying mechanism for the long-term, nonspecific effects of vaccines. J Leukoc Biol 98, 347–56 (2015).

44. Jensen, K.J., Benn, C.S. & van Crevel, R. Unravelling the nature of non-specific effects of vaccines-A challenge for innate immunologists. Semin Immunol 28, 377–83 (2016).

45. Pawlowski, C. et al. Exploratory analysis of immunization records highlights decreased SARS-CoV-2 rates in individuals with recent non-COVID-19 vaccinations. medRxiv, 2020.07.27.20161976 (2020).

46. Debisarun, P.A. et al. The effect of influenza vaccination on trained immunity: impact on COVID-19. medRxiv, 2020.10.14.20212498 (2020).

47. Strope, J.D., Chau, C.H. & Figg, W.D. Are sex discordant outcomes in COVID-19 related to sex hormones? Semin Oncol 47, 335–340 (2020).

48. Mauvais-Jarvis, F., Klein, S.L. & Levin, E.R. Estradiol, Progesterone, Immunomodulation, and COVID-19 Outcomes. Endocrinology 161(2020).

49. Brandi, M.L. & Giustina, A. Sexual Dimorphism of Coronavirus 19 Morbidity and Lethality. Trends Endocrinol Metab 31, 918–927 (2020).

50. Pantos, C., Tseti, I. & Mourouzis, I. Use of triiodothyronine to treat critically ill COVID-19 patients: a new clinical trial. Critical care (London, England) 24, 209–209 (2020).

51. Pantos, C. et al. Triiodothyronine for the treatment of critically ill patients with COVID-19 infection: A structured summary of a study protocol for a randomised controlled trial. Trials 21, 573 (2020).

52. Luxenburger, H. et al. Treatment with proton pump inhibitors increases the risk of secondary infections and ARDS in hospitalized patients with COVID-19: coincidence or underestimated risk factor? Journal of internal medicine, 10.1111/joim.13121 (2020).

53. Kow, C.S. & Hasan, S.S. Use of proton pump inhibitors and risk of adverse clinical outcomes from COVID-19: a meta-analysis. J Intern Med (2020).

54. Flory, C.M. et al. A Preclinical safety study of thyroid hormone instilled into the lungs of healthy rats - an investigational therapy for ARDS. J Pharmacol Exp Ther (2020).

55. Touret, F. et al. In vitro screening of a FDA approved chemical library reveals potential inhibitors of SARS-CoV-2 replication. Sci Rep 10, 13093 (2020).

56. Cadegiani, F.A., McCoy, J., Wambier, C.G. & Goren, A. 5-Alpha-Reductase Inhibitors Reduce Remission Time of COVID-19: Results From a Randomized Double Blind Placebo Controlled Interventional Trial in 130 SARS-CoV-2 Positive Men. medRxiv, 2020.11.16.20232512 (2020).

57. Tan, M.H.E., Li, J., Xu, H.E., Melcher, K. & Yong, E.-l. Androgen receptor: structure, role in prostate cancer and drug discovery. Acta Pharmacologica Sinica 36, 3–23 (2015).

58. Goren, A. et al. Anti-androgens may protect against severe COVID-19 outcomes: results from a prospective cohort study of 77 hospitalized men. Journal of the European Academy of Dermatology and Venereology n/a(2020).

59. Hoffmann, M. et al. SARS-CoV-2 Cell Entry Depends on ACE2 and TMPRSS2 and Is Blocked by a Clinically Proven Protease Inhibitor. Cell 181, 271-280.e8 (2020).

60. Lucas, J.M. et al. The Androgen-Regulated Protease TMPRSS2 Activates a Proteolytic Cascade Involving Components of the Tumor Microenvironment and Promotes Prostate Cancer Metastasis. Cancer Discovery 4, 1310–1325 (2014).

61. Al-Ani, F., Chehade, S. & Lazo-Langner, A. Thrombosis risk associated with COVID-19 infection. A scoping review. Thromb Res 192, 152–160 (2020).

62. Godino, C. et al. Antithrombotic therapy in patients with COVID-19? -Rationale and Evidence. International journal of cardiology, S0167-5273(20)33894-8 (2020).

63. Chow, J.H. et al. Aspirin Use is Associated with Decreased Mechanical Ventilation, ICU Admission, and In-Hospital Mortality in Hospitalized Patients with COVID-19. Anesth Analg (2020).

64. Belsley, D.A. Conditioning diagnostics : collinearity and weak data in regression, (New York (N.Y.) : Wiley, 1991).

65. Neidich, S.D. et al. Increased risk of influenza among vaccinated adults who are obese. International journal of obesity (2005) 41, 1324–1330 (2017).

66. Frasca, D. & Blomberg, B.B. The Impact of Obesity and Metabolic Syndrome on Vaccination Success. Interdiscip Top Gerontol Geriatr 43, 86–97 (2020).

67. Donati Zeppa, S., Agostini, D., Piccoli, G., Stocchi, V. & Sestili, P. Gut Microbiota Status in COVID-19: An Unrecognized Player? Frontiers in Cellular and Infection Microbiology 10(2020).

68. Zuo, T. et al. Alterations in Gut Microbiota of Patients With COVID-19 During Time of Hospitalization. Gastroenterology 159, 944-955.e8 (2020).

69. Vich Vila, A. et al. Impact of commonly used drugs on the composition and metabolic function of the gut microbiota. Nat Commun 11, 362 (2020).

70. McKeigue, P.M. et al. Associations of severe COVID-19 with polypharmacy in the REACT-SCOT case-control study. medRxiv, 2020.07.23.20160747 (2020).

71. Fry, A. et al. Comparison of Sociodemographic and Health-Related Characteristics of UK Biobank Participants With Those of the General Population. American Journal of Epidemiology 186, 1026–1034 (2017).

72. Moskvina, V., Holmans, P., Schmidt, K.M. & Craddock, N. Design of case-controls studies with unscreened controls. Ann Hum Genet 69, 566–76 (2005).

73. Peyrot Wouter J., Boomsma Dorret I., Penninx Brenda W.J.H. & Wray Naomi R. Disease and Polygenic Architecture: Avoid Trio Design and Appropriately Account for Unscreened Control Subjects for Common Disease. The American Journal of Human Genetics 98, 382–391 (2016).

